# The BEYONDD Pilot Study: A Decentralized Community-Engaged Research Framework for Multimodal Characterization of Neurodegenerative Risk in a Multi-Ethnic, Midlife Cohort with Subjective Cognitive or Behavioral Complaints

**DOI:** 10.64898/2026.06.01.26354636

**Authors:** Monica Rivera-Mindt, Micah J. Savin, Vanessa Guzman, Alyssa Arentoft, Eden V. Barragan, Stephen Cubbellotti, Hilary W. Heuer, Kaori Kubo Germano, Howie Rosen, Se Jun Cho, Michelle Higuera, Melissa Sotelo, Chanel Ramirez, Julia E. Culhane, Andrew Margolis, Alexander Slaughter, Amanda Calcetas, Sandra Talavera, Kristen Vincaludo, Joseph DiBenedetto, Omobolanle Ayo, Heining Cham, Gil D. Rabinovici, Adam L. Boxer, Desirée A. Byrd

## Abstract

**Introduction:** Alzheimer’s disease and related dementias (AD/ADRD) pathology begin decades before diagnosis, yet scalable risk detection infrastructures for midlife adults remain limited. The *Biomarker Evaluation of Young Onset Dementia from Diverse Populations* (BEYONDD; R56AG075744) pilot study was designed to address this gap through a decentralized, community-engaged research (CER) model for neurodegenerative risk detection in midlife adults with subjective cognitive or behavioral complaints (sCBC).

**Methods:** This cross-sectional pilot assessed the feasibility of CER-based digital recruitment and participant completion of remotely-acquired screening, cognitive, clinical, and phlebotomy assessments with support of Community Research Navigators (CRNs). Feasibility was evaluated using digital recruitment metrics, yield, retention, and geographic reach.

**Results:** Our approach generated 1.8+ million advertisement impressions, 161,100 clicks, and 4,089 web-registrants. 2,117 individuals completed the online screener, exceeding the prespecified screening goal by 141%. We enrolled a multi-ethnic, midlife cohort of 579 participants (*M_age_*=51.6[6.5]; 75% female; 44% Latinx, 31% non-Latinx Black-American, and 26% all other race/ethnicities), exceeding the enrollment goal by 290%, and 476 participants completed the remote protocol (82% retention). Participants were recruited from 49 U.S. states, Puerto Rico, Australia, and Canada. CRN engagement was concentrated during study stage transitions.

**Discussion:** BEYONDD’s decentralized, CER-based screening infrastructure demonstrated wide geographic reach, strong early-stage engagement, and efficient recruitment among diverse midlife adults. These findings support the feasibility of scalable CER-based digital recruitment for decentralized early detection initiatives and AD/ADRD trials.

## Body

Advances in neurodegenerative plasma biomarkers for Alzheimer’s disease and related dementias (AD/ADRD) have transformed the dementia research landscape, expanding both the methods and temporal windows through which AD/ADRD can be characterized.^1–5^ These advances in AD/ADRD research coincide with a significant demographic reality: ethnocultural populations underrepresented in research (e.g. Black and Latino/a Americans) experience higher prevalence, earlier onset, and greater burden of AD/ADRD.^6–9^ Moreover, these populations are projected to increasingly comprise the majority of AD/ADRD cases in the United States.^10,11^ As a result, emerging biomarker-based risk stratification tools are being calibrated in populations that will not reflect the evolving epidemiologic burden of disease.

Accumulating evidence suggests that midlife molecular changes confer future neurodegenerative risk for AD/ADRD and may represent a critical window for early detection.^12–16^ Midlife detection has implications beyond early-onset AD/ADRD, representing an opportunity for prevention, risk modification, and optimization of treatment response for late-onset disease. ^5,14,17^ However, early neurodegenerative risk detection in midlife adults presents distinct translational challenges, including differences in risk communication, intervention readiness, and etiologic heterogeneity. These challenges are amplified in ethnoculturally underrepresented populations, who experience earlier and greater exposure to modifiable risk factors linked to AD/ADRD and have been profoundly under-included in all current large-scale studies of early onset dementia (EOD), which have lagged behind the emerging demographic diversification accomplished in AD/ADRD studies with older adults.^8,18–21^ As biomarker-based identification shifts earlier in the disease course, representative frameworks for early detection and early onset mild cognitive impairment (EOMCI) and dementia (EOD) diagnosis (e.g., early onset Alzheimer’s disease [EOAD], frontotemporal lobar degeneration [FTLD], Dementia with Lewy Bodies [DLB]) are needed to ensure generalizable translation into timely care and early intervention and prevention trials.

Recent advances in plasma biomarkers have transformed the landscape for early detection in AD/ADRD.^4,22–26^ Blood-based biomarkers are increasingly positioned for use beyond specialty memory clinics, including population-level screening, more real-world community-based settings, and trial enrichment.^4,26^ Emerging research on decentralized models for remote cognitive assessment and biospecimen collection at scale has demonstrated initial promise in cognitively unimpaired, older adult AD/ADRD, and other clinical populations.^27,27–31^ Despite these advances, biomarker studies remain disproportionately focused on older adult (e.g., 65+ years) and primarily homogeneous populations (e.g., ethnocultural status, geography), limiting generalizability and potentially biasing neurodegenerative risk prediction models.^7,32^ The central challenge is therefore no longer biomarker detection, but rather the development and deployment of scalable, decentralized infrastructures for early neurodegenerative risk stratification and validation of these biomarkers in generalizable, midlife adults within community-based settings. Such infrastructures are also needed to improve the repertoire of plasma biomarkers with diagnostic or predictive value in mid-life.

Subjective cognitive and/or behavioral complaints (sCBC) represent one of the earliest phenotypic indicators of AD/ADRD risk and possess potential as a scalable triage signal for early detection of neurodegenerative risk.^33–36^ In older adults, sCBC is associated with amyloid burden, neurodegeneration, and future cognitive decline.^35–37^ In midlife, however, sCBC reflects greater etiologic heterogeneity, ranging from neurodegenerative, psychiatric, and social determinants of health.^9,38,39^ Leveraging sCBC as an initial triage point in midlife, therefore, requires infrastructure that is both biologically anchored and psychosocially informed.

Community-engaged research (CER) models capable of successfully reaching populations historically under-included in AD/ADRD research are essential to address the current gaps in the AD/ADRD early detection research landscape and literature. Recent innovations in applying CER approaches within AD/ADRD studies have demonstrated efficacy in improving demographic representation and the generalizability of this research, including within existing large observational cohorts, such as the Alzheimer’s Disease Neuroimaging Initiative.^40–47^ However, comprehensive deployment of CER strategies has yet to be integrated into decentralized, biomarker characterization approaches for neurodegenerative risk detection in generalizable samples of midlife adults with sCBCs. The *Biomarker Evaluation of Young Onset Dementia from Diverse Populations* (BEYONDD; R56AG075744) is the first nationwide pilot study, of which we are aware, to address these gaps. Specifically, the aim of the BEYONDD pilot study was to utilize CER-based methods to develop and pilot a scalable, decentralized, multimodal phenotyping approach, using *clinically feasible* remote tools (self-report, cognition, & neurodegenerative plasma biomarkers) for early risk detection in a generalizable midlife sCBC sample. Herein, we describe the methods and feasibility of our CER approach within the BEYONDD study.

## METHODS

### Study Design and Setting

The BEYONDD is a cross-sectional cohort building platform and pilot study (R56AG075744) designed to develop and evaluate the feasibility of implementing a large-scale, decentralized approach for midlife neurodegenerative risk detection in adults with sCBC. This study was designed to recruit midlife adults with sCBC outside of specialty memory clinics to characterize early risk in community-based settings.^34^ The pilot phase of BEYONDD was conducted between 2022 and 2026. The study integrated CER-based digital recruitment with remotely-acquired screening, cognitive, clinical, and phlebotomy assessments, and an optional clinic-based validation component (See **Figure 1** for Study Flowchart). All procedures were approved by the Advarra Institutional Review Board (IRB; Columbia, MD, USA) and informed consent was obtained from each participant in accordance with the 1964 Declaration of Helsinki. The present report focuses on BEYONDD’s CER-based methods and feasibility related to our recruitment metrics. Detailed cohort characteristics and clinical findings from the BEYONDD pilot are reported separately.^48^

**Figure 1:**
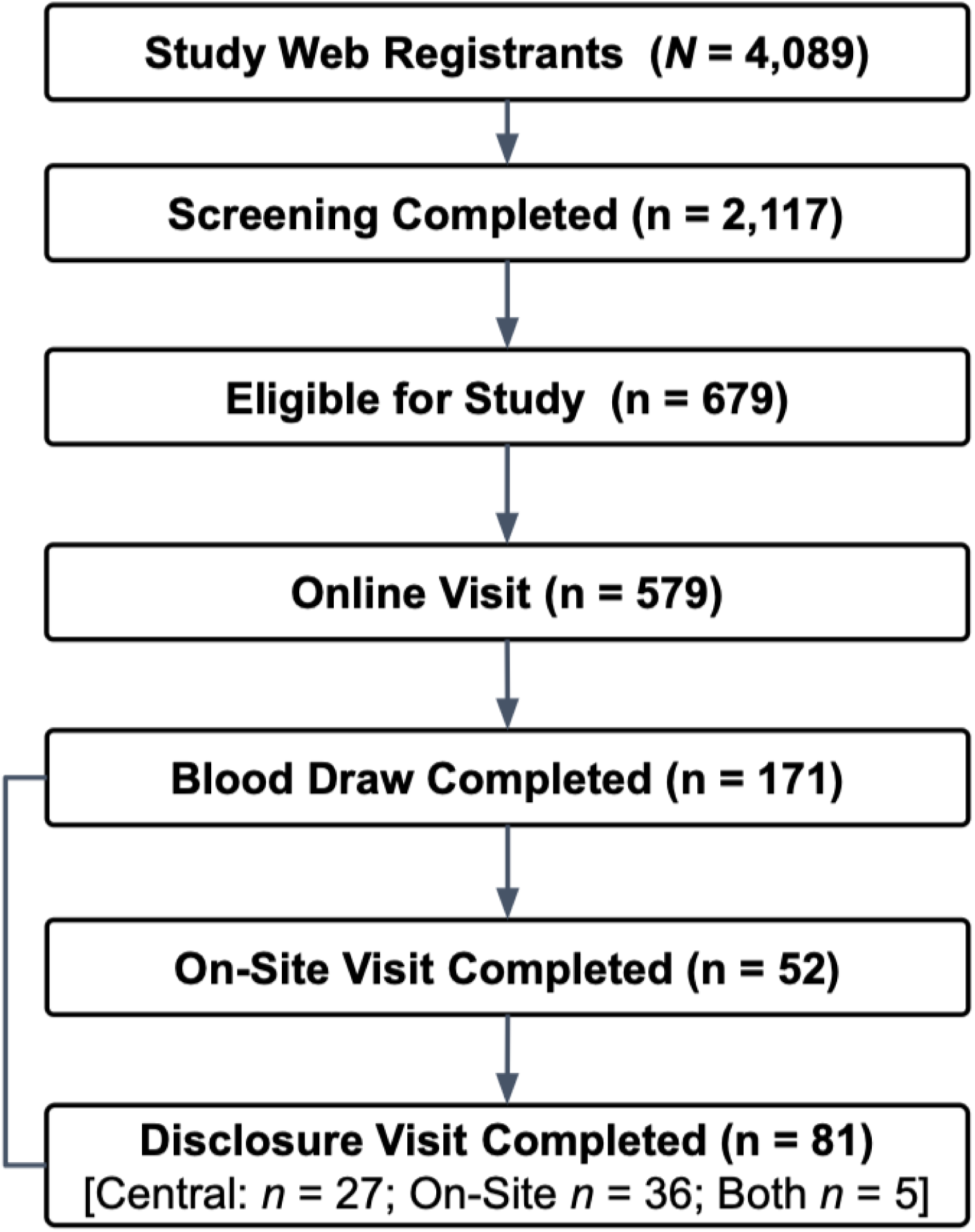
BEYONDD Study Design and Workflow. PRISMA-style flow diagram illustrating participant progression through the Biomarker Evaluation of Young Onset Dementia from Diverse Populations (BEYONDD) study. Participants completed sequential stages including website registration, online screening, eligibility determination, remote assessment, in-home biospecimen collection, optional in-clinic validation, and biomarker results disclosure. The broader BEYONDD infrastructure integrated community-engaged recruitment decentralized multimodal phenotyping, structured return of results, and participant navigation support through Community Research Navigators and the Community-Science Partnership Board.

### Sampling Goals and Inclusion/Exclusion Criteria

Given the profound historical under-inclusion of demographically heterogeneous populations in AD/ADRD research with midlife adults, BEYONDD aimed to oversample community-dwelling adults from ethnoculturally underrepresented populations. To be included in this study, participants needed to be 40 to 64 years of age, identify with an ethnoculturally underrepresented population, endorse subjective cognitive and/or behavioral changes, have access to internet-enabled devices, and be capable of completing digital consent and assessment procedures in either English or Spanish. Individuals with a known diagnosis of dementia, other major neurological disorders, or chronic medical condition related to changes in cognition were excluded.

### Community-Engaged Research Framework for Recruitment

Recruitment and implementation were guided by a structured Community-Engaged Research (CER) framework adapted from Community-Based Participatory Research principles, scaled and adapted for decentralized deployment.^49–51^ For a detailed review of CER elements, please see Rivera Mindt et al., 2024.^40^ In 2022, the BEYONDD Community-Science Partnership Board (CSPB) was established to provide ongoing guidance across study components and met quarterly, offering bidirectional feedback aimed at enhancing cultural responsiveness of the pilot study. The CSPB was comprised of community members, caregivers, and representatives from national AD/ADRD advocacy organizations (see **Acknowledgements** for full membership).

Prior to study launch, recruitment materials were developed in partnership with CSPB members, emphasizing brain health and early detection rather than disease labeling, with attention to minimizing deficit-based language among midlife adults. CSPB contributions also informed the website launch and development of video content for the website, translation and expansion of Spanish-language materials, integration of participant support resources, and creation of referral pathways for individuals not meeting eligibility criteria. In addition, the CSPB provided guidelines on culturally responsive messaging and strategies to improve engagement among underrepresented groups, including approaches used by Community Research Navigators (CRN) who provided remote individualized assistance via phone, email, or text messaging through a HIPAA-compliant communication platform to assist with scheduling, troubleshooting technical issues, and facilitating transitions between study components. CRN support interactions were tracked through a centralized ticketing system that categorized inquiries by study procedure and issue type.

Feedback from the CSPB was incorporated iteratively throughout the study to refine materials and outreach procedures. CSPB consultation contributed to the refinement of recruitment language used in outreach materials, emphasizing accessible terminology and encouraging inclusion of behavioral and speech-related concerns alongside memory-related symptoms in recruitment messaging. CSPB feedback also emphasized the importance of transparent study expectations, responsiveness to participant needs, and enhanced accessibility. As a result, contact information for study staff was more prominently incorporated into recruitment materials and participant communications, and various assessment modalities were made available across study components (e.g., in-home and phone-based). CSPB consultation also emphasized the importance of clear communication, participant autonomy, and referral availability for participants seeking follow-up care. Operational challenges encountered during the pilot phase were presented to CSPB and included scheduling coordination and participant follow-up for in-home visits. Participant navigation and follow-up procedures were iteratively refined with support from CSPB to enhance successful completion of in-home visits in line with approaches described in Rivera Mindt et al., 2024.^40^

### CER-Informed Recruitment

Recruitment was conducted through four complementary engagement pathways: CER-informed digital outreach, community partnerships, clinic and provider referrals, and referrals from related NIH/NIA-funded studies led by BEYONDD investigators (e.g., SALUD [R01AG065110], Black Men’s Brain Health Initiative [R13AG071313]). The primary recruitment approach in BEYONDD was a highly focused CER-informed digital media campaign that was deployed in four U.S. states (California, New York, Texas, and Michigan), Washington, DC, and Puerto Rico, in collaboration with a boutique marketing agency. Recruitment advertisements directed interested individuals to register with the BEYONDD screening portal hosted within the Brain Health Registry.^30^ Media campaign performance was monitored in real time to inform iterative optimization based on cost-per-click, conversion rates, and screener completion yield. Digital recruitment was conducted through paid social media advertising campaigns deployed through the Meta advertising platform. Recruitment campaigns were managed in collaboration with digital media specialists and optimized through an iterative performance monitoring process.

Campaign optimization followed a three-stage approach. First, the initial deployment prioritized broad engagement signals (e.g., advertisement clicks) to identify audience segments interested in study content. Second, campaigns were refined to optimize intermediate engagement metrics such as form completion and study registration. Third, campaign goaling was optimized for qualified participant conversion, defined as enrollment into the remote study protocol. Campaign performance was continuously monitored using platform analytics and study registration metrics. Adjustments to advertisement messaging, imagery, audience goaling, and geographic focus were made iteratively based on observed engagement and conversion performance. Budget allocation was periodically rebalanced to prioritize higher-performing audience segments and geographic regions. Recruitment strategies also included goaled outreach to address demographic imbalances observed during early campaign phases. For example, early campaign stages produced disproportionately higher engagement among women. In response, goaled advertisements aimed at increasing participation among men were introduced despite higher advertising costs associated with demographic goaling. Across successive optimization periods, engagement metrics improved, including increased click-through rates among men in subsequent optimization phases. Campaign messaging, imagery, and geographic goaling were also periodically adjusted based on observed engagement patterns across states, including California, New York, and Texas.

### Screening

After consenting to screening within our online BEYONDD platform within BHR, prospective participants completed a brief online screening battery to assess eligibility, which included questionnaires regarding demographics, medical history, and subjective cognitive and behavioral concerns using the *Cognitive and Behavioral Inventory* (COBI), developed to capture domain-specific self-perceived cognitive and behavioral changes consistent with early phenotypic vulnerability (Barragan et al., in-press). Participants who met eligibility criteria proceeded to electronic consent and enrollment.

### Remote Clinical Assessment

Participants who enrolled in the remote protocol completed an unsupervised, online, brief standardized clinical assessment battery of self-report measures through our secure online platform within BHR (e.g., assessment of demographics; medical and family history; current psychiatric and substance use; sleep; health behaviors; functional status; and social determinants of health). Central CRN staff, located in NYC and San Francisco, provided individualized, remote support for scheduling, troubleshooting, and completion of study procedures. Additional details regarding assessment procedures are described elsewhere (Barragan et al., in-press).

### In-Home Biospecimen Collection

Participants who completed the remote assessment phase were invited to complete in-home phlebotomy through a national mobile phlebotomy service. Biospecimen collection included neurodegenerative plasma biomarkers and routine clinical labs processed using standardized laboratory procedures. Given the logistical complexity of coordinating decentralized in-home visits across states, CRNs provided substantial participant navigation support related to scheduling, follow-up, and visit completion. Detailed biospecimen procedures and assay methods are reported separately (Barragan et al., in-press).

### Remote Cognitive Assessment

Participants additionally completed telephone-based cognitive assessments administered by trained study staff goaling memory, executive functioning, processing speed, and language domains using the National Alzheimer’s Coordinating Center’s Telephone-Administered Cognitive Battery^52^. Optional smartphone-based cognitive and motor assessments were also offered using participants’ personal devices.^19^ Remote cognitive procedures were selected to support decentralized participation and cross-study harmonization approaches used in AD/ADRD research.^53,54^

### In-Clinic Assessment

A subset of participants additionally completed an optional in-person validation visit at seven participating clinical sites across the U.S. (Georgetown University, Washington, DC; Icahn School of Medicine at Mount Sinai, New York, NY; University of California Los Angeles, Los Angeles, CA; University of California San Francisco, San Francisco, CA; University of Michigan, Ann Arbor, MI; University of Texas Health San Antonio, San Antonio, TX) and Puerto Rico (Universidad de Puerto Rico, San Juan, PR). Evaluations included comprehensive neuropsychological assessment, neurological examination, diagnostic consensus procedures using harmonized AD/ADRD research protocols, and optional structural MRI.^55^ Additional details regarding validation procedures are reported separately (Barragan et al., in-press).

### Recruitment Outcomes of Interest

The primary outcomes of this study were selected feasibility- and implementation-related metrics across the decentralized recruitment and enrollment pipeline. Consistent with implementation science frameworks distinguishing feasibility and implementation-related outcomes,^56^ the present study focused on recruitment efficiency, participant engagement, retention, and geographic reach across the AD/ADRD decentralized screening pipeline.^27,28,30^

Feasibility was operationalized using stage-specific yield, conversion rates, and retention indicators across the following study stages: website registration, screening consent, screener completion, eligibility determination, enrollment in the remote study protocol, and remote assessment completion. Stage-specific feasibility benchmarks were established a priori based on projected recruitment capacity, pilot study objectives, and anticipated downstream biomarker and validation procedures, consistent with approaches commonly used in feasibility-oriented cohort development studies.^57,58^ Stage-specific yield was assessed through the extent to which goal goals were met for study registration (*n*=1500), screening (*n*=200), and remote assessments (*n*=200). Conversion rates were calculated at each transition point in the study pipeline, reflecting the proportion of participants who successfully progressed across study stages: registrant-to-screener completion, screener-to-eligibility, eligibility-to-enrollment, and enrollment-to-remote assessment completion. Retention was defined as the proportion of participants who initiated and completed all required procedures within a given study stage: the percent who completed screening once registered, and the percent who completed the remote battery once started. Attrition was defined as the proportion of participants who initiated and discontinued participation within a given stage of the study pipeline. Attrition included participants who were lost to follow-up, declined further participation, or experienced unsuccessful or incomplete study procedures.

Implementation-related metrics included digital recruitment performance, Community Research Navigator engagement, and geographic reach. Digital recruitment performance was evaluated using standard marketing and implementation metrics: total impressions, click-through rates, number of registered participants, and cost-per-click and cost-per-registered participant. Community Research Navigator engagement was evaluated by the proportion of participants who utilized the service. Geographic reach was evaluated by summarizing website registrants’ distribution across U.S. states and Puerto Rico, as well as the number of states in which screening was successfully completed. Representativeness was examined descriptively by comparing participant demographic characteristics to U.S. Census benchmarks for age, sex, race/ethnicity, and educational attainment. Together, these metrics were used to characterize the scalability, engagement, retention, and reach of the BEYONDD decentralized screening infrastructure.

### Statistical Analysis

Recruitment metrics were derived from two primary data sources: (1) advertising performance data from Meta advertising platform analytics, including impressions, clicks, and advertisement engagement metrics, and (2) study pipeline data maintained within the BEYONDD study database, including registration, screening completion, eligibility determination, enrollment, and downstream study procedures. These datasets were integrated to characterize recruitment yield and conversion across stages of the study pipeline. All analyses were conducted using R version 4.5.3. Descriptive statistics were used to summarize digital recruitment performance (impressions, clicks, and registrations), conversion rates across study stages, enrollment into the remote protocol, geographic reach, and completion rates across components. The primary analytic focus was descriptive, summarizing these metrics across predefined stages of the decentralized study pipeline. Continuous variables are reported as means and standard deviations or medians and interquartile ranges, as appropriate, and categorical variables are presented as frequencies and percentages.

## RESULTS

### Overview of Recruitment Pipeline

The decentralized BEYONDD screening infrastructure demonstrated strong engagement across stages of recruitment, screening, and remote assessment (**Figure 1**; **Table 1**). Between January 2023 and January 2025, 4,089 individuals registered through the BEYONDD study website. Of these, 2,559 consented to screening procedures, and 2,117 completed the online eligibility screener. Among participants completing the screener, 679 met eligibility criteria, and 579 enrolled in the remote study protocol. Among enrolled participants, 476 completed the remote psychosocial assessment battery.

**Table 1.** Feasibility, yield, and conversion metrics across the BEYONDD study pipeline. Stage-specific feasibility outcomes across the decentralized Biomarker Evaluation of Young Onset Dementia from Diverse Populations (BEYONDD) study pipeline. Metrics include participant progression from website registration through screening, eligibility determination, enrollment, and remote assessment completion. Conversion rates represent the proportion of participants successfully transitioning between sequential study stages. Prespecified recruitment and assessment benchmarks were used to evaluate feasibility and yield attainment. Overall, findings demonstrate strong feasibility, high participant retention, and successful progression across major stages of the decentralized screening pipeline.

| **Conversion Stages** | **Total Eligible**<br>**(n)** | **Completed**<br>**(n)** | **Conversion Rate**<br>**(%)** |
| --- | --- | --- | --- |
| Screening Consent | 4089 | 2559 | 62.5% |
| Screening Completion | 2559 | 2117 | 82.7% |
| Screening Eligibility | 2117 | 679 | 32.1% |
| Enrollment & Online Visit | 679 | 579 | 85.3% |

### Feasibility

#### Recruitment Goals were Exceeded Resulting in High Yield Attainment

Several prespecified feasibility goals for the BEYONDD pilot were exceeded (See above in **Recruitment Outcome of Interest** for yield goals and rationale). The study aimed to screen approximately 1,500 individuals through digital outreach and online screening procedures. This goal was exceeded by 141%, with 2,117 participants completing the online screener. The pilot also aimed to enroll 200 participants into the remote biomarker screening protocol. This goal was exceeded by 290%, with 579 participants enrolled. Similarly, the study aimed to complete 200 remote psychosocial assessments. This goal was exceeded by 238%, with 476 completed assessments.

### Conversion across and Retention within Study Components were High, with Minimal Attrition

Among 4,089 website registrants, 2,559 consented to screening procedures, corresponding to a 62.5% registrant-to-consent conversion rate. Of those consenting, 2,117 completed the online screener, representing an 82.7% consent-to-screener completion rate.

Attrition during the online screener stage was largely attributable to participants who registered but did not proceed to complete screening procedures. Among the 2,117 individuals completing screening, 679 met eligibility criteria, yielding an eligibility rate of 32.1%. The most common reasons for ineligibility included demographic criteria (*n*=1,130), pre-existing neurological conditions (*n*=223), and absence of subjective cognitive or behavioral complaints (*n*=84).

Among eligible participants, 579 enrolled in the remote study protocol, corresponding to an eligibility-to-enrollment conversion rate of 85.1%. Among participants enrolled in the remote protocol, 476 completed the remote psychosocial assessment battery, corresponding to an 82.2 % enrollment-to-completion yield. Attrition during the remote protocol stage was minimal and reflected incomplete or partially completed remote assessments (17.8 %).

### Implementation Outcomes

#### Digital Recruitment Generated Highly Efficient Engagement

BEYONDD’s digital media campaign generated over 1.8 million total impressions and 528,129 individuals reached, producing 161,100 advertisement clicks, corresponding to a 30.5% click-through rate (**Table 2**). These clicks resulted in 1,917 digital study sign-ups (1.2% click-to-sign-up conversion) and a lower cost per lead of $9.19. Among the 1,917 participants who signed-up with the study, 8.9 % of participants completed blood draws (*n*=171).

**Table 2:** Digital Recruitment Campaign Metrics across the BEYONDD study Pipeline. Digital recruitment funnel metrics describing the BEYONDD decentralized recruitment campaign. Metrics include advertisement impressions, reach, advertisement clicks, digital advertisement sign-ups, cost per Facebook lead, blood draw completion, and downstream clinic referrals. Conversion rate (CVR) represents the percentage of participants progressing from the preceding stage of the digital recruitment funnel. These metrics characterize participant engagement, recruitment efficiency, and progression through major stages of the decentralized study pipeline. Collectively, the findings demonstrate that large-scale digital recruitment can efficiently identify, engage, and advance participants through increasingly intensive stages of neurodegenerative risk characterization.

| Stage | n | CVR % |
| --- | --- | --- |
| Impressions | 1.8M | – |
| Reach | 528,129 | 29.3 % |
| Clicks | 161,100 | 30.5% |
| Digital Ad Sign Ups | 1,917 | 1.2 % |
| Cost per FB Lead | \$9.19 | – |
| Blood Draws | 171 | 8.9% |
| Clinic Referrals | 38 | 29.1% |

### Geographic Reach Exceeded Recruitment Regions

The BEYONDD recruitment campaign surpassed its expectation of recruiting from the 6 base recruitment regions and achieved international reach, with website registrants and participant enrollment occurring in 49 U.S. states, Puerto Rico, with Hawaii representing the only state without registrants (**Figure 2**). At the international level, website registration and participant enrollment occurred in Australia and Canada.

**Figure 2:**
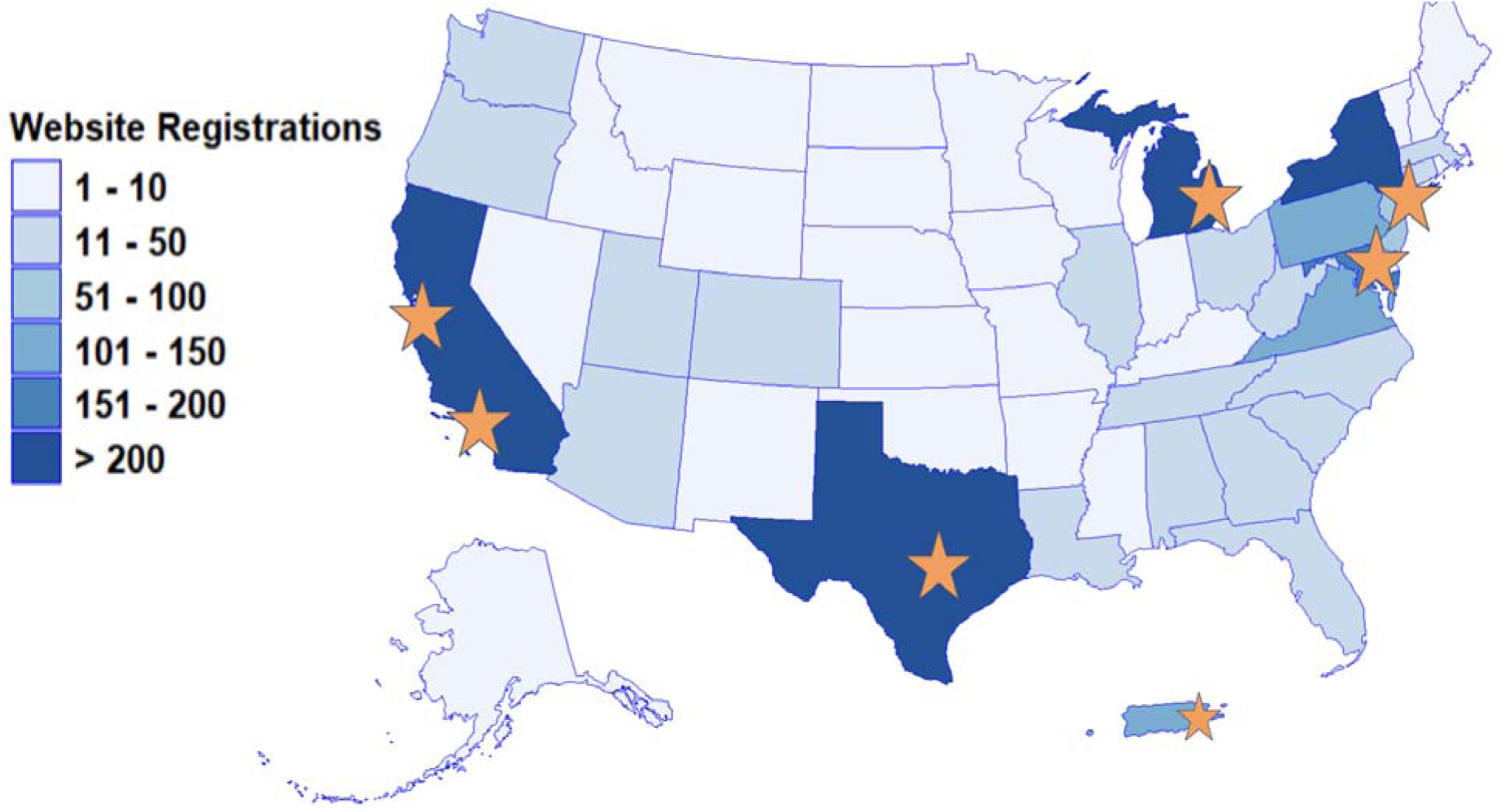
Geographic Reach of Website Registration. Choropleth map illustrating the number of website registrations by state within the Biomarker Evaluation of Young Onset Dementia from Diverse Populations (BEYONDD) study. Blue shading reflects the volume of participant registrations obtained through the decentralized digital recruitment platform, with darker shading indicating greater numbers of registrants. Website registrations were obtained from 49 U.S. states and Puerto Rico, with Hawaii representing the only U.S. state without registrants, demonstrating the broad geographic reach of the BEYONDD recruitment infrastructure. Orange stars denote the seven primary recruitment and validation hub locations: Georgetown University (Washington, DC); Icahn School of Medicine at Mount Sinai (New York, NY); Puerto Rico; University of California, Los Angeles (Los Angeles, CA); University of California, San Francisco (San Francisco, CA); University of Michigan (Ann Arbor, MI); and University of Texas Health San Antonio (San Antonio, TX). Despite recruitment efforts being anchored within these regional hubs, participant enrollment extended well beyond the primary catchment areas, supporting the feasibility of nationwide decentralized neurodegenerative risk screening.

### Community Research Navigators Helped to Bridge the Digital Divide and Supported Complex Logistical Demands

Community Research Navigators (CRNs) provided remote individualized participant support throughout the decentralized screening pipeline. Across the study period, CRNs managed 1,841 remote participant support interactions, including both incoming participant requests and proactive outreach from the study team. Incoming participant inquiries most frequently involved coordination of study visits (*n*=542). Additional inquiries included general study questions (*n*=73) and technical assistance with online assessments (*n*=14).

CRN outreach accounted for a substantial proportion of remote participant interactions (*n*=1,299) and was primarily focused on facilitating completion of study procedures. The largest categories of outgoing CRN interactions involved scheduling and coordinating in-home biospecimen collection visits (*n*=420) and coordinating in-clinic validation visits (*n*=305).

Additional outreach included reminders for questionnaire completion (*n*=127), mobile app visits (*n*=100), and results disclosure sessions (*n*=100). Notably, relatively few support tickets involved technical barriers related to the digital study platform (*n*=7). Instead, the majority of interactions reflected logistical coordination of study procedures, including scheduling, rescheduling, and participant follow-up (*n*=71). Broadly, we observed that CRN engagement was particularly concentrated during transitions between study stages, especially between remote assessment completion and scheduling of in-home biospecimen collection visits.

### Participant Characteristics were Highly Diverse, Educated, and Primarily Female

Participant demographic characteristics are summarized in **Figure 3**. Following completion of the online visit, the cohort *(n=*579) represented a true midlife sample, with a mean age of 52 years (*SD*=6.5; range=41–64 years), and was predominantly female (75.4%). The cohort demonstrated substantial ethnocultural diversity, including 44% Latino/a participants, 31% non-Latino Black/African American participants, 10% Asian participants, 9% multiracial participants, and 6% identifying as other or unknown race/ethnicity. Educational attainment was broad, ranging from less than a high school diploma to postgraduate education, with the majority of participants completing at least some college education (*M* years of education=15.5, *SD*=4). Compared descriptively with U.S. Census benchmarks, the BEYONDD cohort demonstrated greater representation of populations historically underrepresented in AD/ADRD research, consistent with the study’s community-engaged recruitment framework.

**Figure 3:**
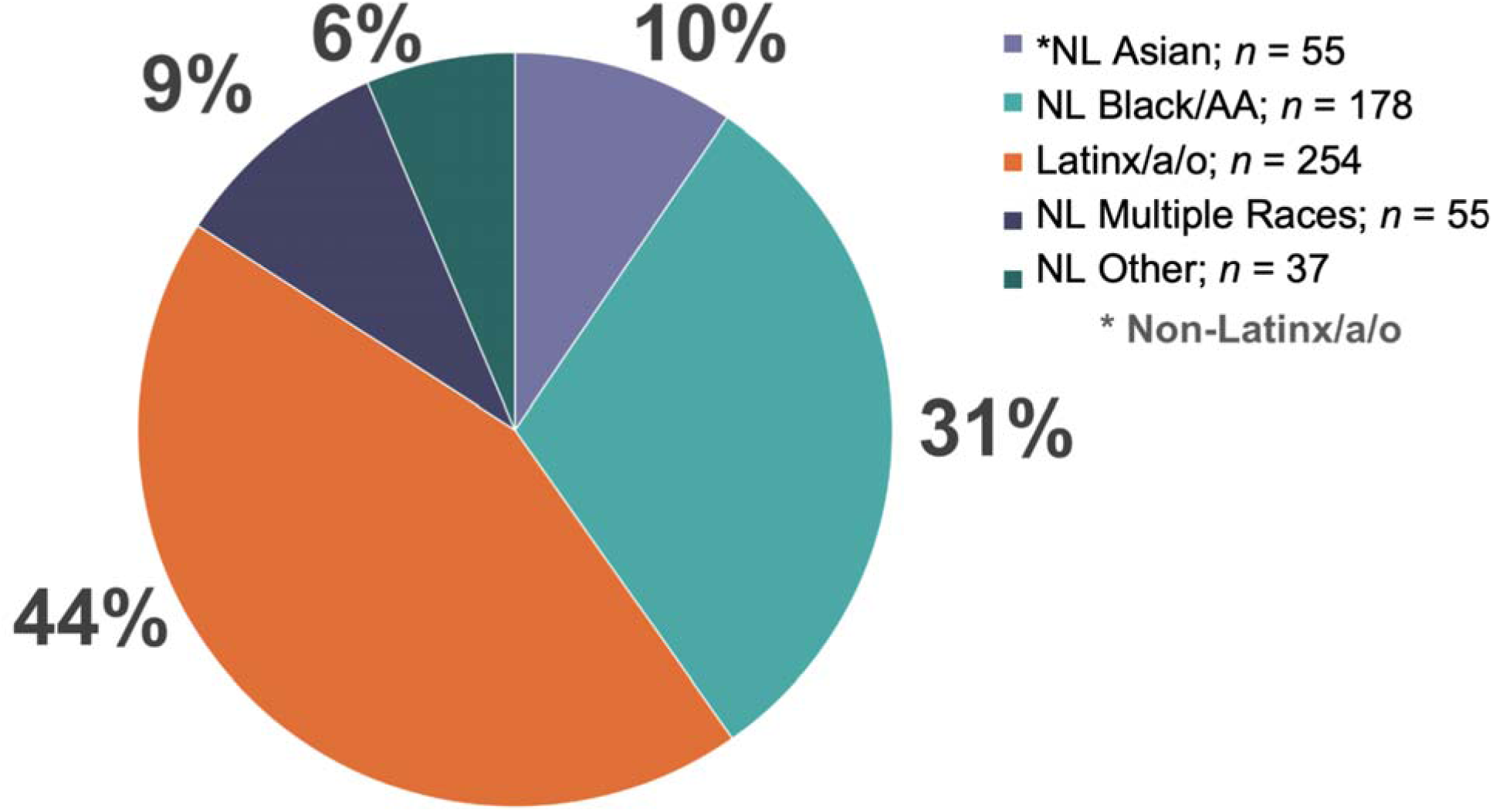
Ethnocultural Composition of Participants Enrolled in the BEYONDD Cohort. Pie chart illustrating the self-reported race and ethnicity of participants who completed the BEYONDD remote assessment visit (n = 579). The cohort was predominantly composed of participants from ethnoculturally underrepresented populations, including Latinx/a/o, Black/African American, Asian, and multiracial individuals. Values represent participant counts (n) and percentages (%) within each category. NL = Non-Latinx/a/o; AA = African American.

## DISCUSSION

The present findings provide proof-of-concept support for what is, to our knowledge, the first nationwide decentralized, community-engaged, multimodal phenotyping infrastructure designed to characterize neurodegenerative risk in a diverse cohort of midlife adults with sCBC. BEYONDD successfully recruited a community-based midlife cohort, with participants spanning ages 40 – 64 years and a mean age of 52 years, thereby addressing a major gap within the current AD/ADRD research landscape, which remains disproportionately centered on older adult and specialty clinic populations.^12–16,53^ The study additionally exceeded multiple prespecified recruitment and engagement benchmarks, including screening, enrollment, and remote assessment completion goals, while achieving broad geographic reach across 49 U.S. states, Puerto Rico, Australia, and Canada. The cohort also demonstrated substantially greater representation of ethnoculturally underrepresented populations relative to many existing biomarker and observational cohorts, with ∼85% of the cohort being comprised of participants from Latinx, Black-American, or Asian populations. Collectively, these findings suggest that decentralized, CER-informed infrastructures integrating digital recruitment, remote participant navigation, and remote multimodal phenotyping may provide a scalable and transportable framework for early neurodegenerative risk characterization in representative midlife populations. These findings have implications for screening infrastructure, biomarker interpretation, and AD/ADRD prevention research.

From an infrastructure development perspective, BEYONDD highlights an emerging methodological challenge within biomarker-era AD/ADRD research: scientific advances in plasma biomarkers, remote assessment technologies, and prevention science are rapidly outpacing the development of representative infrastructures capable of deploying these advances within heterogeneous community populations.^1–5,22–27,27,29,30^ Existing observational cohorts and prevention-oriented studies have substantially advanced understanding of AD/ADRD biology, yet many remain anchored to recruitment models centered within specialty clinics, older adult populations, and geographically concentrated academic medical centers.^28–30,40,42–44,59,60^ Consequently, populations experiencing disproportionate AD/ADRD burden remain underrepresented in neurodegenerative plasma biomarker studies, prevention trials, and early-stage observational cohorts.^6–9,61^ As biomarker-based identification increasingly shifts toward earlier stages of disease detection, the central translational challenge is no longer solely biomarker discovery, but rather the development of representative and transportable infrastructures capable of identifying neurodegenerative vulnerability prior to specialty clinic presentation and substantial disease progression.

From a scientific perspective, the AD/ADRD transportability challenge is not solely logistical, but interpretive. Neurodegenerative plasma biomarker meaning is shaped by the populations within which risk is characterized, the pathways through which individuals enter research, and the broader clinical and social contexts in which phenotypic vulnerability emerges. Midlife adults with sCBC represent a particularly heterogeneous population in whom neurodegenerative risk may intersect with psychiatric symptoms, vascular burden, chronic stress exposure, social determinants of health, and barriers to healthcare access. Existing biomarker cohorts have often characterized disease after substantial progression and following specialty clinic ascertainment, potentially narrowing the range of clinical and social heterogeneity captured within biomarker-defined populations. BEYONDD was intentionally designed to address this translational gap through community-based ascertainment, decentralized multimodal phenotyping, and participant-centered engagement strategies capable of identifying neurodegenerative vulnerability earlier and within heterogeneous populations, and community-based settings. Notably, this approach enabled inclusion of community-dwelling midlife adults from Puerto Rico with subjective cognitive and behavioral complaints, representing, to our knowledge, the first decentralized neurodegenerative risk and plasma biomarker study in this population. BEYONDD thereby reached groups historically underrepresented in ADRD biomarker research despite barriers to specialty neurological care.^62,63^ These findings suggest that future AD/ADRD prevention infrastructures may need to be designed around scalable community-based vulnerability characterization rather than specialty clinic ascertainment alone.

In response to these scientific and implementation challenges, BEYONDD was i designed not simply as a screening study or observational cohort, but as a decentralized infrastructure for early neurodegenerative risk characterization embedded within real-world community settings. The study integrated CER-informed recruitment, adaptive digital engagement strategies, remote cognitive and biomarker phenotyping, participant navigation systems, and optional clinic-based harmonized validation procedures within a single scalable framework. In doing so, BEYONDD advances an alternative model for biomarker-era AD/ADRD infrastructure development, one that prioritizes representativeness, accessibility, scalability, and biological characterization. This distinction may be important for midlife populations, in whom subjective cognitive and behavioral concerns emerge within highly heterogeneous and socially embedded contexts that are often not captured within conventional clinic-based ascertainment models.

A key methodological feature of the BEYONDD infrastructure was the integration of community-engaged research (CER) principles throughout study development, implementation, and recruitment procedures. Community engagement within BEYONDD functioned not simply as an outreach strategy, but as a methodological component of infrastructure development.

Recruitment materials, participant support systems, and study procedures were developed iteratively in collaboration with the Community-Science Partnership Board (CSPB), composed of community members, caregivers, and representatives from national advocacy organizations.

CSPB guidance informed culturally responsive communication practices, participant-centered engagement strategies, and approaches designed to minimize deficit-oriented framing of cognitive concerns among midlife adults.^40,42–46,49,50,60^ These processes were particularly important given longstanding mistrust of biomedical research and persistent barriers to participation among populations historically underrepresented in AD/ADRD research.^42–46,49,50,64^ While prior studies have demonstrated the importance of community engagement for improving participation in AD/ADRD research,^40,42–46,49,50,60^ the present findings extend this literature by illustrating how CER frameworks can be operationalized within decentralized biomarker-era infrastructures to support trust, engagement, and sustained participation across geographically distributed populations.

Consistent with CER evidence, the study demonstrated strong engagement and recruitment efficiency across the decentralized study pipeline. Digital recruitment campaigns generated over 1.8 million advertisement impressions and 161,000 advertisement clicks, ultimately producing enrollment and remote assessment completion rates that substantially exceeded prespecified feasibility benchmarks. These recruitment strategies incorporated iterative optimization informed by real-time engagement analytics, allowing recruitment messaging and tailored responsivity to evolve dynamically across implementation phases.

Rather than functioning as isolated digital marketing approaches, these adaptive recruitment strategies were embedded within a broader CER-informed framework designed to improve representativeness and engagement among populations historically under-included in AD/ADRD research.^28–30,40,51,59,60^ Together, these findings suggest that scalable digital recruitment approaches, when integrated within participant-centered and culturally responsive infrastructures, may represent an important pathway toward developing more representative biomarker-era cohorts.

Beyond recruitment itself, the present findings highlight the critical role of Community Research Navigators (CRNs) within decentralized multimodal research infrastructures. CRNs provided remote, individualized support across study transitions, including scheduling, follow-up, troubleshooting, and coordination of increasingly complex procedures such as in-home biospecimen collection and clinic-based validation visits. Importantly, CRN engagement was particularly concentrated during transitions to later-stage study components, where logistical complexity and participant burden increased substantially. Notably, relatively few participant interactions involved technological barriers, whereas the majority reflected logistical coordination demands associated with sustaining engagement across multi-stage decentralized participation. These observations suggest that successful decentralized AD/ADRD infrastructures may depend less upon technology alone and more upon participant-centered navigation systems capable of supporting remote continuity of engagement across increasingly complex biomarker and phenotyping procedures.^28,30,42,59,64^

Beyond implementation outcomes, the recruitment findings carry important implications for the representativeness and transportability of biomarker-era AD/ADRD research.

Populations historically underrepresented in AD/ADRD research experience disproportionate burden of dementia yet remain substantially under-included in biomarker studies and prevention trials.^6–9,61^ Structural barriers—including geographic distance from specialty centers, limited access to dementia-focused care, and longstanding mistrust of biomedical research—continue to shape participation within observational cohorts and clinical trials.^42,43,64,65^ The BEYONDD framework was intentionally designed to address several of these barriers through decentralized participation pathways, bilingual engagement procedures, and CER-informed recruitment strategies. As prevention-oriented trials increasingly move toward earlier stages of disease detection, representative decentralized cohorts will become increasingly essential not only for equitable participation, but also for understanding how biomarker-defined vulnerability manifests across heterogeneous populations and lived contexts.^4,26,28,29,66^

Finally, the present findings support midlife as a critical methodological window for neurodegenerative risk characterization and prevention science. Neurodegenerative processes associated with AD/ADRD begin decades prior to clinical diagnosis, creating a prolonged preclinical interval during which early intervention may be most effective.^12–16,53^ Despite this, many existing observational cohorts and biomarker studies continue to recruit primarily from older adult populations or specialty clinic settings after substantial disease progression has already occurred.^28,31^ BEYONDD advances an alternative infrastructure model in which subjective cognitive and behavioral complaints function as an early phenotypic entry point for scalable neurodegenerative vulnerability characterization within community settings.^33–39^ While sCBC reflects heterogeneous etiologies within midlife populations, pairing these concerns with decentralized biomarker, cognitive, and clinical phenotyping approaches may provide a pragmatic pathway for identifying individuals appropriate for downstream characterization, monitoring, and prevention-oriented intervention studies.^22–26,32–39^ Future work will be needed to determine how these multidimensional phenotypic and biomarker profiles relate to longitudinal trajectories of cognitive decline, resilience, and treatment responsiveness across diverse populations.

Several limitations should be considered when interpreting these findings. First, the present report focused primarily on feasibility- and implementation-related outcomes rather than biological or longitudinal clinical findings. Downstream biomarker analyses and clinic-based validation outcomes from the BEYONDD cohort will be reported separately. Second, participation required internet-enabled devices and sufficient digital literacy to complete remote procedures, which may introduce selection biases despite the decentralized design. Finally, while the intentional focus on ethnoculturally underrepresented populations may limit direct comparison with historically homogeneous cohorts, this design reflects the broader scientific objective of improving representativeness within biomarker-era AD/ADRD research infrastructures.

In conclusion, the BEYONDD is the first nationwide pilot study to provide empirical support for a novel decentralized, community-engaged infrastructure capable of supporting scalable neurodegenerative vulnerability characterization among diverse midlife adults with sCBC. The findings demonstrate that CER methodology, digital recruitment science, participant navigation systems, and decentralized multimodal phenotyping can be successfully integrated within a single transportable framework to support the next generation of prevention-oriented AD/ADRD research. As blood-based biomarkers and early intervention approaches continue to shift the field toward earlier stages of disease detection, the future utility of these technologies will depend not only upon analytic validity, but upon the representativeness, scalability, and contextual grounding of the infrastructures through which neurodegenerative vulnerability is characterized.^1–5,26,29^

## Data Availability

All data produced in the present study are available upon reasonable request to the authors.

## ACKNOWLEDGEMENTS

The authors gratefully acknowledge the contributions of BEYONDD participants and study partners; the BEYONDD Community-Science Partnership Board (CSPB), including community stakeholders, caregivers, and representatives from national AD/ADRD advocacy organizations who provided ongoing consultation throughout study development and implementation. CSPB members included Angela Donadelle, Anita Bhama, Ashley Jackson, Brandon Feldt, Sandra Gonzalez-Morett, and representatives from our industry partners: Keith Fargo and Robin Otto for the Lewy Body Dementia Association; Rebecca Edelmayer and Percy Griffin for the Alzheimer’s Association; Shana Dodge, Penny Dacks, and Sharon Denny for the Association for Frontotemporal Degeneration; Lakecia Vincent and Carrie Milliard for the Frontotemporal Degeneration Disorders Registry; our seven BEYONDD site leaders and teams (Georgetown University, Washington, DC [PI: Raymond Scott Turner]; Icahn School of Medicine at Mount Sinai, New York, NY; University of California Los Angeles, Los Angeles [PI: Maryam Belgi], CA; University of California San Francisco, San Francisco, CA; University of Michigan, Ann Arbor, MI [PI: Navid Seraji-Bozorgzad]; University of Texas Health San Antonio, San Antonio, TX [PI: Campbell Sullivan]); and the Rivera Mindt Lab Spanish Capacity Work Group for their leadership in translating BEYONDD materials to Spanish. We also wish to extend our deep gratitude to the Alzheimer’s Association (Maria Carrillo, Carl Hill, and Heather Snyder) for their early and sustained support in the development and deployment of the BEYONDD pilot study. The authors additionally acknowledge the contributions of Alaniz Marketing, including Roxanne Alaniz, Jennefer Sorce, and John Olin for their support with community-engaged digital recruitment strategies. The authors are grateful for the contributions of our past team members Adeyinka Ajayi and Annabelle Soto.

## Author Contributions

MRM, DB, ALB, and GR conceptualized the research design. All authors contributed to drafting, critical revision, and final approval of the manuscript.

## Ethics Statement

The BEYONDD study was approved by the Institutional Review Boards of participating institutions. All procedures were conducted in accordance with the Declaration of Helsinki and relevant federal guidelines for human subjects research. All participants provided electronic informed consent prior to participation. Additional consent was obtained for in-home biospecimen collection and optional in-clinic evaluations. Procedures for biomarker disclosure included clinical oversight and systematic monitoring for adverse events.

## References

1. Jack CR, Knopman DS, Jagust WJ, et al. Hypothetical model of dynamic biomarkers of the Alzheimer’s pathological cascade. Lancet Neurol. 2010;9(1):119–128. doi:10.1016/S1474-4422(09)70299-6

2. Jack CR, Bennett DA, Blennow K, et al. NIA-AA Research Framework: Toward a biological definition of Alzheimer’s disease. Alzheimers Dement J Alzheimers Assoc. 2018;14(4):535–562. doi:10.1016/j.jalz.2018.02.018

3. Sperling RA, Aisen PS, Beckett LA, et al. Toward defining the preclinical stages of Alzheimer’s disease: recommendations from the National Institute on Aging-Alzheimer’s Association workgroups on diagnostic guidelines for Alzheimer’s disease. Alzheimers Dement J Alzheimers Assoc. 2011;7(3):280–292. doi:10.1016/j.jalz.2011.03.003

4. Hansson O, Edelmayer RM, Boxer AL, et al. The Alzheimer’s Association appropriate use recommendations for blood biomarkers in Alzheimer’s disease. Alzheimers Dement J Alzheimers Assoc. 2022;18(12):2669–2686. doi:10.1002/alz.12756

5. Jack Jr. CR, Andrews JS, Beach TG, et al. Revised criteria for diagnosis and staging of Alzheimer’s disease: Alzheimer’s Association Workgroup. Alzheimers Dement. 2024;20(8):5143–5169. doi:10.1002/alz.13859

6. Adkins-Jackson PB, George KM, Besser LM, et al. The structural and social determinants of Alzheimer’s disease related dementias. Alzheimers Dement J Alzheimers Assoc. 2023;19(7):3171–3185. doi:10.1002/alz.13027

7. Raman R, Quiroz YT, Langford O, et al. Disparities by Race and Ethnicity Among Adults Recruited for a Preclinical Alzheimer Disease Trial. JAMA Netw Open. 2021;4(7):e2114364. doi:10.1001/jamanetworkopen.2021.14364

8. Mayeda ER, Glymour MM, Quesenberry CP, Whitmer RA. Inequalities in dementia incidence between six racial and ethnic groups over 14 years. Alzheimers Dement J Alzheimers Assoc. 2016;12(3):216–224. doi:10.1016/j.jalz.2015.12.007

9. Nianogo RA, Rosenwohl-Mack A, Yaffe K, Carrasco A, Hoffmann CM, Barnes DE. Risk Factors Associated With Alzheimer Disease and Related Dementias by Sex and Race and Ethnicity in the US. JAMA Neurol. 2022;79(6):584–591. doi:10.1001/jamaneurol.2022.0976

10. Matthews KA, Xu W, Gaglioti AH, et al. Racial and ethnic estimates of Alzheimer’s disease and related dementias in the United States (2015-2060) in adults aged ≥65 years. Alzheimers Dement J Alzheimers Assoc. 2019;15(1):17–24. doi:10.1016/j.jalz.2018.06.3063

11. Alzheimer’s Association 2024 Alzheimer’s Disease Facts and Figures.

12. Bateman RJ, Xiong C, Benzinger TLS, et al. Clinical and biomarker changes in dominantly inherited Alzheimer’s disease. N Engl J Med. 2012;367(9):795–804. doi:10.1056/NEJMoa1202753

13. Villemagne VL, Burnham S, Bourgeat P, et al. Amyloid β deposition, neurodegeneration, and cognitive decline in sporadic Alzheimer’s disease: a prospective cohort study. Lancet Neurol. 2013;12(4):357–367. doi:10.1016/S1474-4422(13)70044-9

14. Livingston G, Huntley J, Sommerlad A, et al. Dementia prevention, intervention, and care: 2020 report of the Lancet Commission. Lancet. 2020;396(10248):413–446. doi:10.1016/S0140-6736(20)30367-6

15. Lane CA, Hardy J, Schott JM. Alzheimer’s disease. Eur J Neurol. 2018;25(1):59-70. doi:10.1111/ene.13439

16. Iturria-Medina Y, Sotero RC, Toussaint PJ, Mateos-Pérez JM, Evans AC, Alzheimer’s Disease Neuroimaging Initiative. Early role of vascular dysregulation on late-onset Alzheimer’s disease based on multifactorial data-driven analysis. Nat Commun. 2016;7:11934. doi:10.1038/ncomms11934

17. Dubois B, Villain N, Frisoni GB, et al. Clinical diagnosis of Alzheimer’s disease: recommendations of the International Working Group. Lancet Neurol. 2021;20(6):484–496. doi:10.1016/S1474-4422(21)00066-1

18. Apostolova LG, Aisen P, Eloyan A, et al. The Longitudinal Early-onset Alzheimer’s Disease Study (LEADS): Framework and methodology. Alzheimers Dement J Alzheimers Assoc. 2021;17(12):2043–2055. doi:10.1002/alz.12350

19. Paolillo EW, Casaletto KB, Clark AL, et al. Examining Associations Between Smartphone Use and Clinical Severity in Frontotemporal Dementia: Proof-of-Concept Study. JMIR Aging. 2024;7(1):e52831. doi:10.2196/52831

20. Mukadam N, Marston L, Lewis G, et al. South Asian, Black and White ethnicity and the effect of potentially modifiable risk factors for dementia: A study in English electronic health records. PLOS ONE. 2023;18(10):e0289893. doi:10.1371/journal.pone.0289893

21. Mindt MR, Okonkwo O, Weiner MW, et al. Improving generalizability and study design of Alzheimer’s disease cohort studies in the United States by including under-represented populations. Alzheimers Dement J Alzheimers Assoc. 2023;19(4):1549–1557. doi:10.1002/alz.12823

22. Palmqvist S, Janelidze S, Quiroz YT, et al. Discriminative Accuracy of Plasma Phospho-tau217 for Alzheimer Disease vs Other Neurodegenerative Disorders. JAMA. 2020;324(8):772–781. doi:10.1001/jama.2020.12134

23. Karikari TK, Pascoal TA, Ashton NJ, et al. Blood phosphorylated tau 181 as a biomarker for Alzheimer’s disease: a diagnostic performance and prediction modelling study using data from four prospective cohorts. Lancet Neurol. 2020;19(5):422–433. doi:10.1016/S1474-4422(20)30071-5

24. Palmqvist S, Warmenhoven N, Anastasi F, et al. Plasma phospho-tau217 for Alzheimer’s disease diagnosis in primary and secondary care using a fully automated platform. Nat Med. 2025;31(6):2036–2043. doi:10.1038/s41591-025-03622-w

25. Moscoso A, Grothe MJ, Ashton NJ, et al. Time course of phosphorylated-tau181 in blood across the Alzheimer’s disease spectrum. Brain J Neurol. 2021;144(1):325–339. doi:10.1093/brain/awaa399

26. Duncan GB, Dickson SP, Kaplan JM, et al. Leveraging recent advances in plasma biomarkers to optimize early proof of concept trials in Alzheimer’s disease. Alzheimers Dement Transl Res Clin Interv. 2025;11(4):e70183. doi:10.1002/trc2.70183

27. Walter S, Langford O, Jimenez-Maggiora GA, et al. The AlzMatch Pilot Study - Feasibility of Remote Blood Collection of Plasma Biomarkers for Preclinical Alzheimer’s Disease Trials. J Prev Alzheimers Dis. 2024;11(5):1435–1444. doi:10.14283/jpad.2024.101

28. Walter S, Langford OG, Clanton TB, et al. The Trial-Ready Cohort for Preclinical and Prodromal Alzheimer’s Disease (TRC-PAD): Experience from the First 3 Years. J Prev Alzheimers Dis. 2020;7(4):234–241. doi:10.14283/jpad.2020.47

29. Rafii MS, Sperling RA, Donohue MC, et al. The AHEAD 3-45 Study: Design of a prevention trial for Alzheimer’s disease. Alzheimers Dement. 2023;19(4):1227–1233. doi:10.1002/alz.12748

30. Weiner MW, Nosheny R, Camacho M, et al. The Brain Health Registry: An internet-based platform for recruitment, assessment, and longitudinal monitoring of participants for neuroscience studies. Alzheimers Dement J Alzheimers Assoc. 2018;14(8):1063–1076. doi:10.1016/j.jalz.2018.02.021

31. DuBord AY, Paolillo EW, Staffaroni AM. Remote Digital Technologies for the Early Detection and Monitoring of Cognitive Decline in Patients With Type 2 Diabetes: Insights From Studies of Neurodegenerative Diseases. J Diabetes Sci Technol. 2024;18(6):1489–1499. doi:10.1177/19322968231171399

32. Mm M, Jl D, Rd F, et al. Performance of plasma phosphorylated tau 181 and 217 in the community. Nat Med. 2022;28(7). doi:10.1038/s41591-022-01822-2

33. Jessen F, Amariglio RE, van Boxtel M, et al. A conceptual framework for research on subjective cognitive decline in preclinical Alzheimer’s disease. Alzheimers Dement J Alzheimers Assoc. 2014;10(6):844–852. doi:10.1016/j.jalz.2014.01.001

34. Rabin LA, Smart CM, Amariglio RE. Subjective Cognitive Decline in Preclinical Alzheimer’s Disease. Annu Rev Clin Psychol. 2017;13:369–396. doi:10.1146/annurev-clinpsy-032816-045136

35. Buckley RF, Maruff P, Ames D, et al. Subjective memory decline predicts greater rates of clinical progression in preclinical Alzheimer’s disease. Alzheimers Dement J Alzheimers Assoc. 2016;12(7):796–804. doi:10.1016/j.jalz.2015.12.013

36. Nosheny RL, Amariglio R, Sikkes SAM, et al. The role of dyadic cognitive report and subjective cognitive decline in early ADRD clinical research and trials: Current knowledge, gaps, and recommendations. Alzheimers Dement Transl Res Clin Interv. 2022;8(1):e12357. doi:10.1002/trc2.12357

37. Amariglio RE, Becker JA, Carmasin J, et al. Subjective cognitive complaints and amyloid burden in cognitively normal older individuals. Neuropsychologia. 2012;50(12):2880–2886. doi:10.1016/j.neuropsychologia.2012.08.011

38. Hill NL, Mogle J, Wion R, et al. Subjective Cognitive Impairment and Affective Symptoms: A Systematic Review. The Gerontologist. 2016;56(6):e109–e127. doi:10.1093/geront/gnw091

39. Parfenov VA, Zakharov VV, Kabaeva AR, Vakhnina NV. Subjective cognitive decline as a predictor of future cognitive decline: a systematic review. Dement Neuropsychol. 2020;14(3):248–257. doi:10.1590/1980-57642020dn14-030007

40. Rivera Mindt M, Arentoft A, Calcetas AT, et al. The Alzheimer’s Disease Neuroimaging Initiative-4 (ADNI-4) Engagement Core: A culturally informed, community-engaged research (CI-CER) model to advance brain health equity. Alzheimers Dement. 2024;20(12):8279–8293. doi:10.1002/alz.14242

41. Ashford MT, Raman R, Miller G, et al. Screening and enrollment of underrepresented ethnocultural and educational populations in the Alzheimer’s Disease Neuroimaging Initiative (ADNI). Alzheimers Dement J Alzheimers Assoc. 2022;18(12):2603–2613. doi:10.1002/alz.12640

42. Gilmore-Bykovskyi AL, Jin Y, Gleason C, et al. Recruitment and retention of underrepresented populations in Alzheimer’s disease research: A systematic review. Alzheimers Dement. 2019;5:751–770. doi:10.1016/j.trci.2019.09.018

43. Glover CM, Shah RC, Bennett DA, Wilson RS, Barnes LL. The Health Equity Through Aging Research And Discussion (HEARD) Study: A Proposed Two-Phase Sequential Mixed-Methods Research Design To Understand Barriers And Facilitators Of Brain Donation Among Diverse Older Adults. Exp Aging Res. 2020;46(4):311–322. doi:10.1080/0361073X.2020.1747266

44. Flatt JD, Stites SD, Kittle KR, Cicero EC, Anderson JG, Wharton W. Promoting Inclusion of Sexual and Gender Minority Individuals in Aging and Alzheimer’s Disease and Related Dementias Research. Alzheimers Dement. 2022;18(S11):e062324. doi:10.1002/alz.062324

45. Perales-Puchalt J, Shaw A, McGee JL, et al. Preliminary Efficacy of a Recruitment Educational Strategy on Alzheimer’s Disease Knowledge, Research Participation Attitudes, and Enrollment Among Hispanics. Hisp Health Care Int Off J Natl Assoc Hisp Nurses. 2020;18(3):144–149. doi:10.1177/1540415319893238

46. Maestre GE, Blangero J, Manusov EG, et al. Decentralizing dementia research to the US-Mexico border: the Rio Grande Valley AD-RCMAR as a model for translational equity. Alzheimers Dement. 2026;12(2):e70238. doi:10.1002/trc2.70238

47. Green-Harris G, Coley SL, Koscik RL, et al. Addressing Disparities in Alzheimer’s Disease and African-American Participation in Research: An Asset-Based Community Development Approach. Front Aging Neurosci. 2019;11. doi:10.3389/fnagi.2019.00125

48. Alzheimer’s Association International Conference. ALZ - Alzheimer’s Association International Conference. Accessed May 29, 2026. https://alz.confex.com/alz/2024/meetingapp.cgi/Index/CB_Topic∼Human

49. Israel BA, Schulz AJ, Parker EA, Becker AB. Review of community-based research: assessing partnership approaches to improve public health. Annu Rev Public Health. 1998;19:173–202. doi:10.1146/annurev.publhealth.19.1.173

50. Wallerstein N, Duran B. Community-Based Participatory Research Contributions to Intervention Research: The Intersection of Science and Practice to Improve Health Equity. Am J Public Health. 2010;100(Suppl 1):S40–S46. doi:10.2105/AJPH.2009.184036

51. Ashford MT, Camacho MR, Jin C, et al. Digital culturally tailored marketing for enrolling Latino participants in a web-based registry: Baseline metrics from the Brain Health Registry. Alzheimers Dement J Alzheimers Assoc. 2023;19(5):1714–1728. doi:10.1002/alz.12805

52. 52. Gierzynski TF, Gregoire A, Reader JM, et al. Evaluation of the Uniform Data Set version 3 teleneuropsychological measures. J Int Neuropsychol Soc JINS. 2024;30(2):183-193. doi:10.1017/S1355617723000383

53. Dubois B, Villain N, Frisoni GB, et al. Clinical diagnosis of Alzheimer’s disease: recommendations of the International Working Group. Lancet Neurol. 2021;20(6):484–496. doi:10.1016/S1474-4422(21)00066-1

54. 54. Weintraub S, Besser L, Dodge HH, et al. Version 3 of the Alzheimer Disease Centers’ Neuropsychological Test Battery in the Uniform Data Set (UDS). Alzheimer Dis Assoc Disord. 2018;32(1):10-17. doi:10.1097/WAD.0000000000000223

55. 55. Weintraub S, Salmon D, Mercaldo N, et al. The Alzheimer’s Disease Centers’ Uniform Data Set (UDS): the neuropsychologic test battery. Alzheimer Dis Assoc Disord. 2009;23(2):91-101. doi:10.1097/WAD.0b013e318191c7dd

56. Proctor E, Silmere H, Raghavan R, et al. Outcomes for implementation research: conceptual distinctions, measurement challenges, and research agenda. Adm Policy Ment Health. 2011;38(2):65–76. doi:10.1007/s10488-010-0319-7

57. Leon AC, Davis LL, Kraemer HC. The role and interpretation of pilot studies in clinical research. J Psychiatr Res. 2011;45(5):626–629. doi:10.1016/j.jpsychires.2010.10.008

58. Billingham SAM, Whitehead AL, Julious SA. An audit of sample sizes for pilot and feasibility trials being undertaken in the United Kingdom registered in the United Kingdom Clinical Research Network database. BMC Med Res Methodol. 2013;13:104. doi:10.1186/1471-2288-13-104

59. Walter S, Langford O, Jimenez-Maggiora GA, et al. The AlzMatch Pilot Study - Feasibility of Remote Blood Collection of Plasma Biomarkers for Preclinical Alzheimer’s Disease Trials. J Prev Alzheimers Dis. 2024;11(5):1435–1444. doi:10.14283/jpad.2024.101

60. Ashford MT, Raman R, Miller G, et al. Screening and enrollment of underrepresented ethnocultural and educational populations in the Alzheimer’s Disease Neuroimaging Initiative (ADNI). Alzheimers Dement. 2022;18(12):2603–2613. doi:10.1002/alz.12640

61. Matthews KA, Xu W, Gaglioti AH, et al. Racial and ethnic estimates of Alzheimer’s disease and related dementias in the United States (2015-2060) in adults aged ≥65 years. Alzheimers Dement J Alzheimers Assoc. 2019;15(1):17–24. doi:10.1016/j.jalz.2018.06.3063

62. Buckley T. RELATIONSHIP BETWEEN PHYSICAL AND MENTAL HEALTH WITH SUBJECTIVE COGNITIVE DECLINE AMONG OLDER ADULTS IN PUERTO RICO. Innov Aging. 2022;6(Suppl 1):672-673. doi:10.1093/geroni/igac059.2475

63. Olivari B, Taylor C, McGuire L. Improving Usefulness of Cognitive Decline Population Measures in Predicting Future Dementia Burden. Innov Aging. 2021;5(Suppl 1):181–182. doi:10.1093/geroni/igab046.692

64. George S, Duran N, Norris K. A systematic review of barriers and facilitators to minority research participation among African Americans, Latinos, Asian Americans, and Pacific Islanders. Am J Public Health. 2014;104(2):e16–31. doi:10.2105/AJPH.2013.301706

65. Denny A, Streitz M, Stock K, et al. Perspective on the “African American participation in Alzheimer disease research: Effective strategies” workshop, 2018. Alzheimers Dement J Alzheimers Assoc. 2020;16(12):1734-1744. doi:10.1002/alz.12160

66. Ritchie M, Hussen K, Langford O, et al. Recruitment and retention in a preclinical AD trial: comparisons between academic and non-academic sites. Alzheimers Res Ther. 2025;17(1):222. doi:10.1186/s13195-025-01867-8

